# Lead exposure and serum metabolite profiles in pregnant women in Mexico City

**DOI:** 10.1101/2021.06.03.21258309

**Authors:** Megan M. Niedzwiecki, Shoshannah Eggers, Anu Joshi, Georgia Dolios, Alejandra Cantoral, Héctor Lamadrid-Figueroa, Chitra Amarasiriwardena, Martha M. Téllez-Rojo, Robert O. Wright, Lauren Petrick

## Abstract

**Background:** Lead (Pb) exposure is a global health hazard causing a wide range of adverse health outcomes. Yet, the mechanisms of Pb toxicology remain incompletely understood, especially during pregnancy. To uncover biological pathways impacted by Pb exposure, this study investigated serum metabolomic profiles during the third trimester of pregnancy that are associated with blood Pb and bone Pb.

**Methods:** We used data and specimens collected from 99 women enrolled in the Programming Research in Obesity, Growth, Environment, and Social Stressors birth cohort based in Mexico City. Maternal Pb exposure was measured in whole blood samples from the third trimester of pregnancy and in the tibia and patella bones at 1 month postpartum. Third-trimester serum samples underwent metabolomic analysis; metabolites were identified based on matching to an in-house analytical standard library. A metabolome-wide association study was performed with all three Pb measurements using multiple linear regression models, adjusted for confounders and batch effects. Class enrichment analyses were also conducted.

**Results:** The median (interquartile range) blood Pb concentration was 2.9 (2.6) μg/dL. Median bone Pb, measured in the patella and tibia, were 2.5 (7.3) μg/g and 3.6 (9.5) μg/g, respectively. Of 248 total metabolites identified in serum, 31 were associated with blood Pb (p<0.05). Class enrichment analysis identified significant overrepresentation of metabolites classified as fatty acids and conjugates, amino acids and peptides, and purines. Tibia and patella Pb were associated with 14 and 11 metabolites, respectively (p<0.05). Comparing results from bone and blood Pb, glycochenodeoxycholic acid and glycocholic acid were negatively associated with blood Pb and tibia Pb, while 5-aminopentanoic acid and 7-methylguanine were negatively associated with blood Pb and patella Pb. One metabolite, 5-aminopentanoic acid, was associated with all three Pb measures.

**Conclusion:** This study identified serum metabolites in pregnant women associated with Pb measured in blood (31 metabolites) and bone (tibia: 14 metabolites, patella: 11 metabolites). These findings provide insights on the metabolic profile around Pb exposure in pregnancy and may provide important links to guide detailed studies of toxicological effects for both mothers and children.

## Background

Lead (Pb) exposure is a persistent global health hazard, with no safe exposure threshold (1). Pb is a naturally occurring non-essential element, and human industrial practices can promote Pb exposures through the contamination of dust, food, and water (2). Once ingested, Pb often substitutes for calcium (Ca^2+^) in the human body and can accumulate in bone, where it is stored long-term and can re-enter the bloodstream during periods of bone remodeling (3). Pb exposures are linked to a wide array of detrimental health outcomes (2,4,5), including neurotoxicity, reduced kidney function, anemia, joint weakness, hypertension, reduced immune function, and reproductive complications.

Children are particularly susceptible to the effects of Pb exposures: for a given Pb exposure, the relative exposure in children is larger by body mass than in adults, and children have higher soft tissue Pb absorption (3,6,7). Further, children have a more permeable blood-brain barrier than adults, making the neurotoxic effects of Pb particularly pronounced (3). Finally, there are multiple critical neurodevelopmental windows during childhood wherein Pb exposure may lead to life-long adverse neurological effects (8,9). Despite these significant health concerns, the mechanisms of Pb toxicity are not fully understood. Novel approaches to investigating Pb metabolism and toxicology may promote new avenues for discovering these mechanisms. Additionally, further examination of Pb exposure during pregnancy is particularly crucial, as exposure during this period can exert harmful effects in both mothers and children.

One powerful approach to uncover the biological impacts of Pb exposure is through metabolomics. Measuring metabolites in biological samples, such as plasma or serum, provides a snapshot of metabolic function, enabling the investigation of biological responses to exposures, including Pb. Unlike other ‘omic technologies (e.g., genomics, transcriptomics, etc.) that capture the *potential* for phenotypic changes, metabolomics shows the *actual* state of metabolic function. Therefore, metabolomics can reveal the metabolic links between Pb exposure and downstream health outcomes by investigating metabolites and pathways strongly associated with Pb exposure.

Metabolomics analyses of Pb exposures are relatively sparse. The few metabolomics studies of Pb have primarily been conducted in animals, adult males, and highly-exposed populations. In a study investigating the effects of Pb exposure on the gut microbiome and blood metabolome in 8-week-old female mice, Gao et al. found that Pb exposure altered multiple metabolomic pathways including nitrogen metabolism, oxidative stress, vitamin E, and bile acids (10). A similar dosing study in 8-month-old rats found that Pb exposure reduced levels of butyryl-L-carnitine and ganglioside GD2 (d18:0/20:0) and increased levels of metabolites associated with oxidative stress pathways (11). In an epidemiologic investigation of the urinary metabolome of highly-exposed participants residing near a Pb acid battery recycling plant, blood Pb was associated with several metabolic pathways, including porphyrin, amino acid, and chlorophyll metabolism, heme biosynthesis, and ATP-binding cassette transporters (12). Another epidemiologic analysis in the Veterans Administration Normative Aging Study found several blood metabolites associated with blood Pb, including amino acids, lipids, and metabolites involved in oxidative stress and immune pathways (13). Taken together, these results indicate that biologically-relevant metabolic changes occur with Pb exposure. While some findings are consistent (amino acids, oxidative stress), numerous factors, including the timing, level, and population(s) of Pb exposure, may affect metabolomic responses. Because pregnancy is a uniquely important period of the lifecourse, understanding Pb-associated metabolomic changes in pregnant women–particularly in response to low-level Pb exposures in ranges commonly observed in human populations–is critical to broadly understand the impacts of Pb exposure on human health.

In observational studies, Pb can be measured in a variety of samples–including hair, nails, urine, blood, and bone–with each matrix capturing a different window of exposure. Blood and bone, which are considered the gold standards for Pb measurement, capture short- and long-term exposure, respectively: Pb has a half-life from 1 week to 2 months in blood, depending on age and exposure history, and 5 to 20 years in bone (14). Previous metabolomics studies primarily measured Pb in blood, which captures relatively recent Pb exposure. Metabolomic analyses in relation to bone Pb will add important information to the literature on the biological impacts of longer-term Pb exposure.

This study investigated serum metabolomic profiles during the third trimester of pregnancy to provide insights on the biological impacts of short- and long-term Pb exposures during this critically important period. We profiled metabolites and measured Pb levels in third-trimester blood samples and collected postpartum bone Pb measures for 99 women in the Programming Research in Obesity, Growth, Environment, and Social Stressors (PROGRESS) cohort based in Mexico City, Mexico.

## Methods

### Study subjects

The PROGRESS cohort recruited women who were pregnant and receiving prenatal care through the Mexican Social Security System (Instituto Mexicano del Seguro Social – IMSS) between July 2007 and February 2011; in total, there were 948 women with live births. Women were eligible to participate in the study if they were ≥18 years old and <20 weeks of gestation, planned to stay in Mexico City for the next 3 years, had access to a telephone, had no medical history of heart or kidney disease, did not consume alcohol daily, and did not use any steroid or anti-epilepsy medications. Procedures were approved by institutional review boards at the Harvard School of Public Health, Icahn School of Medicine at Mount Sinai, and the Mexican National Institute of Public Health. Women provided written informed consent in Spanish. For the current study, 99 women with available serum samples stored at −80^°^C from the third trimester of pregnancy were randomly selected from the full cohort for metabolomics analysis.

### Lead (Pb) measurements

All Pb measurements were previously analyzed as described (15). In short, blood specimens in trace metal-free tubes were analyzed using an Agilent 8800 ICP Triple Quad in MS/MS mode in the trace metals laboratory at the Icahn School of Medicine at Mount Sinai. The limit of detection was <0.2 μg/dL, and the instrument precision was approximately 5% relative standard deviation. At 1 month postpartum, mother’s tibia (cortical bone) and patella (trabecular bone) Pb concentrations were measured using a K-shell X-ray fluorescence instrument, a noninvasive, low-radiation-dose method of measuring bone lead content (16). We estimated lead concentration for 30 min for each leg and the measures were averaged by the inverse of the proportion of the measurement error corresponding to each determination. Bone Pb content is thought to provide an indicator of exposure over the span of decades; in particular, tibia measurements reflect longer time spans (>10 years) compared to patella (1-5 years) (17).

### Metabolomics analysis

Stored serum samples were thawed on ice and vortexed, and 50-uL aliquots were transferred to a microcentrifuge tube. 150 uL of methanol containing internal standards was added, and the sample vortexed then incubated at −20°C for 30 min. Samples were centrifuged, the supernatant dried using a Savant SC250EXP SpeedVac concentrator at 35°C for 90 minutes and stored at −80°C until analysis. Before liquid chromatography high-resolution mass spectrometry (LC-HRMS) analysis, dried extracts were reconstituted in 60-uL of 80% acetonitrile or 100% methanol for analysis using zwitterionic hydrophilic interaction liquid chromatography (ZIC-HILIC) or reverse phase (RP) LC connected to HRMS in positive and negative modes, respectively, as described elsewhere (18,19). An additional 10-uL aliquot from each sample was combined for use as a pooled quality control sample (“pooled QC”) and processed similarly. Following the same protocol, matrix blanks (replacing the plasma with water) and multiple pooled QCs were extracted. Samples were analyzed in a randomized order in four analytical batches, with pooled QCs injected routinely throughout the run.

Metabolites were identified based upon in-house database matching considering retention time, accurate mass, and MS/MS matching (when available) with pure standards analyzed under the same conditions. Peaks were identified and integrated using the Personal Chemical Database Library and Profinder software (Agilent Technologies, Santa Clara, USA). There were 139 metabolites and 115 metabolites detected by HILIC and RP chromatography methods, respectively. Of these, seven metabolites were identified using both methods; only the higher intensity value was retained for analyses. Therefore, the combined dataset had 248 metabolites of interest. Metabolites were evaluated for missing values and only retained if they had more than 50% non-missing values. Missing values were imputed with the minimum value of the metabolite divided by the square root of 2. Imputed data were then log_2_-transformed to remove heteroscedasticity (unequal variability) in the metabolite intensities.

### Covariates

Age and educational attainment were ascertained at enrollment. Since only one woman in the cohort reported smoking tobacco during pregnancy, environmental tobacco smoke exposure during pregnancy was characterized as the report of any tobacco smoker in the home during the second or third trimester of pregnancy. Body mass index (BMI) was determined using height (in meters [m]) and weight (in kilograms [kg]) ascertained at the third-trimester study visit. The season of the last menstrual period was defined according to weather patterns in Mexico City as dry/cold (January-February; November-December), dry/warm (March-April) and rainy (May-October). Since metabolite levels can vary by laboratory batch, models also accounted for the metabolomics batch assignment as a categorical variable.

### Statistical analysis

A metabolome-wide association study approach was used to identify metabolites associated with blood and bone Pb measures. Using this approach, multiple linear regression models were constructed for each Pb variable (blood Pb, tibia Pb, or patella Pb) as the main predictor of log_2_-transformed metabolite intensities, adjusting for all covariates listed above. For significant findings, the patterns of the associations were examined using bivariate scatterplots and locally estimated scatterplot smoothing regression. For bone Pb variables, negative instrument values were included in analyses; sensitivity analyses were conducted in which negative values were imputed with simulated values between 0 and the detection limit (2 μg/g) generated from a uniform distribution.

To identify significant metabolites while accounting for the high correlation among the metabolite measurements, we implemented the “number of effective tests” adjustment approach previously used in metabolomics studies (13,20,21). Briefly, we used principal components analysis to identify the number of principal components (PCs) that explain 75%, 95%, and 99% of the variance in metabolite intensities, then used these numbers to calculate adjusted p-values (*a*/*m*), where *a* reflects the nominal p-value threshold of 0.05, and *m* reflects the number of PCs that explain the defined level of variance. Herein, we calculated effective number of test (ENT) thresholds of 75% (ENT75%: 0.05/24=0.002083), 95% (ENT95%: 0.05/60=0.00083), and 99% (ENT99%: 0.05/84=0.000595).

For biological interpretation of the blood Pb results, we input metabolites into MetaboAnalyst (22) to perform class- and pathway-based enrichment analyses. We identified significant metabolite classes and metabolic pathways using the main-class chemical structure and the Small Molecule Pathway Database (23) pathway-based metabolite set libraries, respectively. For co-eluting metabolites, only the first listed metabolite was used as input in enrichment analyses.

## Results

The study sample consisted of 99 randomly-selected pregnant women from the PROGRESS cohort with complete data on exposure and covariates (**Table 1**). The median (interquartile range [IQR]) age of these women was 27.7 (6.8) years, and 63% completed a high school education or greater.

**Table 1.** Descriptive characteristics for PROGRESS mothers in current study (N=99).

| **Characteristics** | **Level/Unit** | **Median (IQR) or N (%)** |
| --- | --- | --- |
| ***Demographics*** |  |  |
| **Age at recruitment** | Years | 27.7 (6.8) |
| **BMI in 3<sup>rd</sup> trimester** | kg/m<sup>2</sup> | 28.6 (5.0) |
| **Maternal Education** | < High School | 37 (37%) |
|  | High School | 38 (38%) |
|  | > High School | 24 (24%) |
| **Smoke exposure** | Yes | 30 (30%) |
| **Season of last menstrual period** | Dry/cold | 29 (29%) |
|  | Dry/warm | 22 (22%) |
|  | Rainy | 48 (48%) |
| ***Lead (Pb) exposure*** |  |  |
| **Blood Pb, 3rd trimester** | µg/dL | 2.9 (2.6) |
| **Tibia Pb, 1 month postpartum** | µg/g | 2.5 (7.3) |
| **Patella Pb, 1 month postpartum** | µg/g | 3.6 (9.5) |

The median (IQR) Pb concentration in third-trimester blood samples was 2.9 (2.6) μg/dL (**Table 1**). These levels are higher than those observed in non-pregnant women of childbearing age measured in NHANES during a similar time period (24). Current guidelines for medical management of lead-exposed adults recommend that pregnant women avoid occupations and avocational exposures that may result in blood Pb levels > 5 μg/dL (25). In our study, nineteen mothers had blood Pb concentrations above the CDC action level of ≥ 5 μg/dL for pregnant women; among these women, three had Pb levels of concern (≥ 10 μg/dL) (26).

Bone patella and tibia Pb levels measured at 1 month postpartum—reflecting long-term Pb exposures from 1–5 years and >10 years, respectively—were 2.5 (7.3) μg/g and 3.6 (9.5) μg/g, respectively (**Table 1**). The Pb levels in patella and tibia were strongly correlated (Pearson’s r=0.51), while, as expected, bone Pb measures were less strongly correlated with blood Pb (patella and blood Pb, r=0.34; tibia and blood Pb, r=0.15) (**Figure 1**).

**Figure 1.**
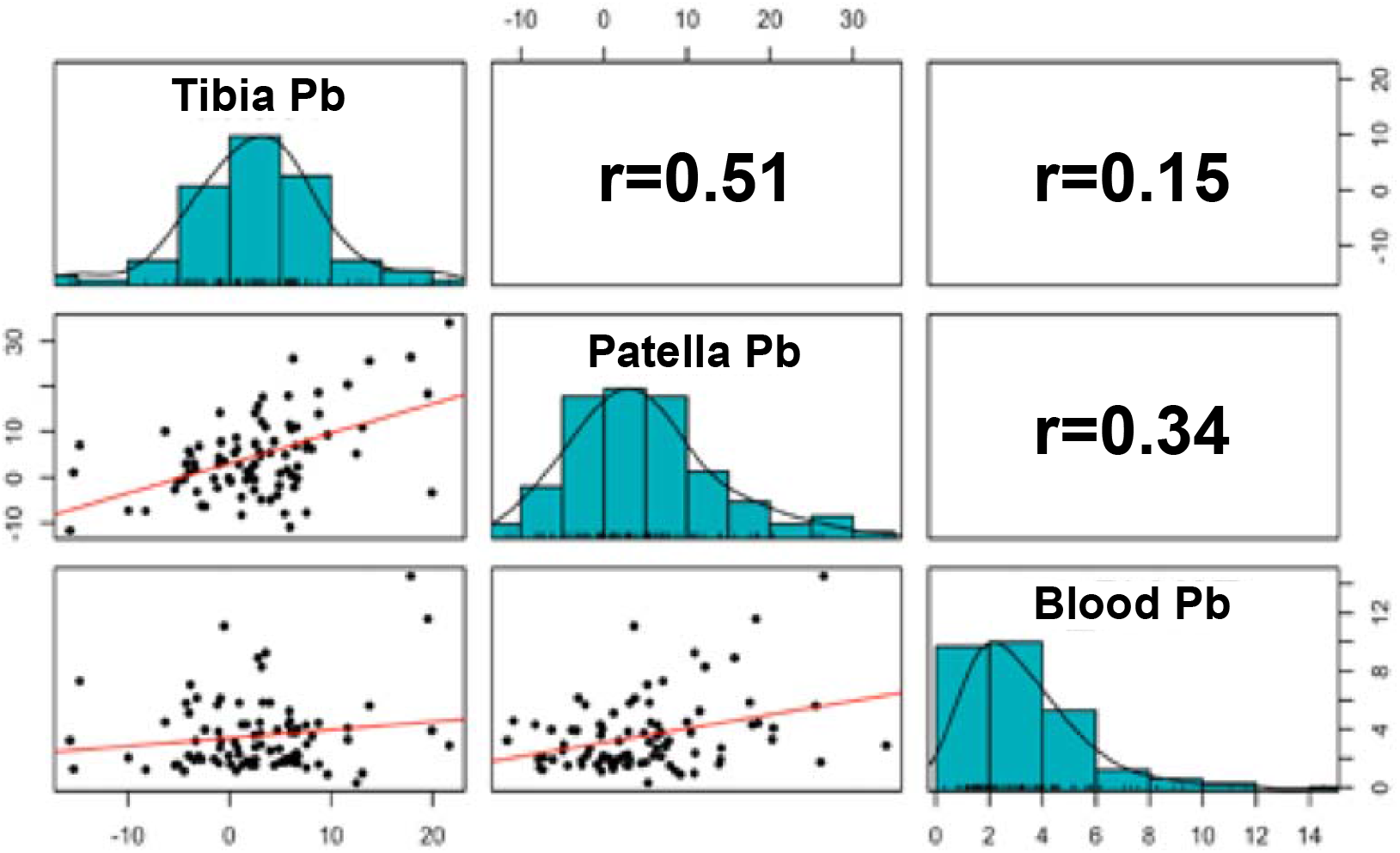
Distributions and correlations of Pb in 3rd trimester blood, tibia, and patella.

We detected a total of 248 metabolites in third-trimester serum samples. Of these, 31 metabolites were associated with third-trimester blood Pb at nominal significance (p<0.05) (**Table 2**), and 2-hydroxybutyrate and hypoxanthine was negatively associated at ENT95% and ENT75% thresholds, respectively. Class-based enrichment analyses of these 31 metabolites identified significant overrepresentation (Holm-Bonferroni p<0.05) of metabolites classified as fatty acids and conjugates, amino acids and peptides, and purines (noted with asterisks in **Table 2**).

**Table 2.** Serum metabolites associated with blood Pb in the 3^rd^ trimester.

| Metabolite subclass | Compound<sup>a</sup> | Estimate (SE) | P |
| --- | --- | --- | --- |
| Fatty acids and conjugates*** | Myristic acid | -0.073 (0.024) | 2.63E-03 |
|  | Stearic acid | -0.043 (0.014) | 3.02E-03 |
|  | Adrenic acid | -0.058 (0.019) | 3.37E-03 |
|  | Palmitoleic acid | -0.097 (0.032) | 3.56E-03 |
|  | Oleic acid / Elaidic acid / Petroselinic acid | -0.051 (0.020) | 1.25E-02 |
|  | Arachidonic acid | -0.040 (0.016) | 1.52E-02 |
|  | Lauric acid | -0.058 (0.023) | 1.53E-02 |
|  | Dihomo-Gamma-Linolenic acid | -0.040 (0.016) | 1.69E-02 |
|  | Margaric acid / Hexadecanol | -0.046 (0.020) | 2.14E-02 |
|  | Arachidic acid | -0.036 (0.016) | 2.91E-02 |
|  | Palmitic acid | -0.041 (0.014) | 5.14E-03 |
|  | Eicosapentaenoic acid | -0.049 (0.024) | 4.40E-02 |
| Amino acids, peptides, and analogues*** | N-Acetylasparagine | 0.152 (0.052) | 4.09E-03 |
|  | 5-Aminopentanoate | -0.137 (0.053) | 1.16E-02 |
|  | N-Acetylalanine | -0.047 (0.019) | 1.48E-02 |
|  | Asparagine | -0.035 (0.016) | 3.05E-02 |
|  | N-Acetylmethionine | -0.084 (0.040) | 3.94E-02 |
|  | Creatine | -0.064 (0.031) | 4.00E-02 |
|  | L-Leucine | -0.039 (0.019) | 4.16E-02 |
| Purines and purine derivatives*** | Hypoxanthine | -0.047 (0.014) | 1.26E-03 |
|  | 7-Methylguanine | -0.015 (0.006) | 1.70E-02 |
|  | Caffeine | 0.193 (0.080) | 1.84E-02 |
|  | 1-Methyluric acid | 0.131 (0.062) | 3.85E-02 |
| Bile acids, alcohols and derivatives | Glycocholic acid | 0.079 (0.035) | 2.73E-02 |
|  | Glycochenodeoxycholic acid | 0.075 (0.038) | 4.93E-02 |
| Alpha hydroxy acids and derivatives | 2-Hydroxybutyrate | -0.055 (0.016) | 6.50E-04 |
| Dicarboxylic acids and derivatives | Succinate | -0.062 (0.024) | 1.17E-02 |
| Organosulfonic acids and derivatives | Taurine | -0.037 (0.015) | 1.26E-02 |
| Benzoic acids and derivatives | MBP (Mono-n-butyl phthalate) | -0.076 (0.036) | 3.85E-02 |
| Linoleic acids and derivatives | Alpha-linolenic acid (ALA) / Gamma-Linolenic acid (GLA) | -0.079 (0.032) | 1.50E-02 |
| Fatty acid esters | Acetylcarnitine | -0.049 (0.017) | 4.16E-03 |

\*\*\*Significantly-enriched (Holm-Bonferroni $p < 0.05$ ) subclass from enrichment analysis in MetaboAnalyst; a. Compounds that co-eluted and could not be discriminated indicated with “/”

Additionally, pathway-based enrichment analysis identified the alpha linolenic acid and linoleic acid metabolic pathway (**Figure 2**), which contained five metabolites that were negatively associated with blood Pb. We note that linoleic acid and the *n*-3 fatty acid docosahexaenoic acid (DHA), the latter of which is synthesized in this pathway, were also negatively associated with blood Pb at marginal significance (linoleic acid: estimate [SE], −0.043 [0.022], p=0.06; DHA: estimate [SE], −0.038 [0.020], p=0.06).

**Figure 2.**
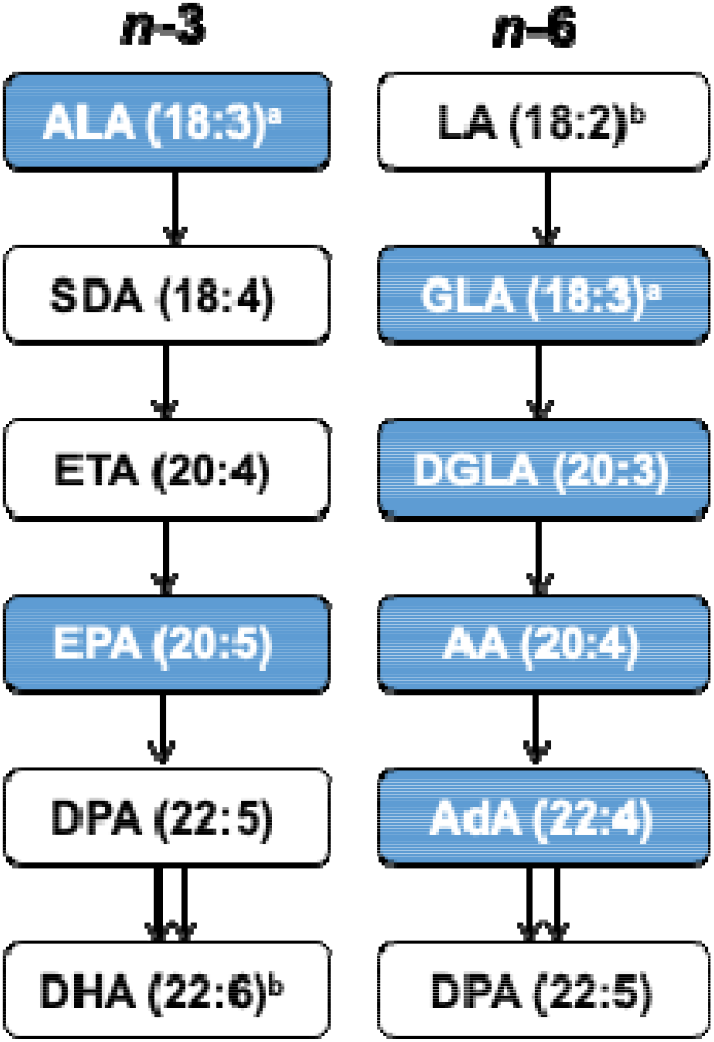
Pathway enrichment analysis of serum metabolites associated with blood Pb identifies the alpha linolenic acid and linoleic acid pathway. Metabolites negatively associated with blood Pb (p<0.05) levels shown in blue. ^a^ALA and GLA co-eluted; ^b^LA and DHA were negatively associated with blood Pb at marginal significance (p=0.06). ALA: alpha linolenic acid; SDA: stearidonic acid; ETA: eicosatetraenoic acid; EPA: eicosapentaenoic acid; DPA: docosapentaenoic acid; DHA: docosahexaenoic acid; LA: linoleic acid; GLA: gamma-linolenic acid; DGLA: dihomo-gamma-linolenic acid; AA: arachidonic acid; AdA: adrenic acid.

We also examined third-trimester serum metabolites associated with patella and tibia Pb, which identified 14 and 11 metabolites, respectively, at p<0.05 (**Table 3**); of these, two —5-aminopentanoic acid and L-arginine—were negatively associated with both bone Pb measures. Betaine was positively associated with patella Pb at ENT99%, and 1-linoleoylglycerol was positively associated with tibia Pb at ENT95%. In sensitivity analyses, results did not materially change when negative instrument values for bone Pb were imputed with positive values below the limit of detection (data not shown).

**Table 3.** Serum metabolites associated with patella and tibia Pb.

|  | Patella Pb |  | Tibia Pb |  |
| --- | --- | --- | --- | --- |
| Compound | Estimate (SE) | P | Estimate (SE) | P |
| *Significant in patella and tibia* |  |  |  |  |
| 5-Aminopentanoic acid | -0.049 (0.015) | 1.82E-03 | -0.049 (0.022) | 2.46E-02 |
| L-Arginine | -0.010 (0.005) | 4.70E-02 | -0.015 (0.007) | 2.74E-02 |
| *Significant in patella* |  |  |  |  |
| Betaine | 0.031 (0.009) | 5.31E-04 | 0.018 (0.014) | 0.19 |
| Corticosterone | -0.102 (0.039) | 1.12E-02 | -0.089 (0.056) | 0.11 |
| Biliverdin | 0.017 (0.007) | 1.97E-02 | 0.007 (0.010) | 0.45 |
| Oxoproline | 0.012 (0.005) | 2.05E-02 | 0.004 (0.007) | 0.60 |
| Pyroglutamate | 0.012 (0.005) | 2.09E-02 | 0.004 (0.007) | 0.60 |
| Lithocholic acid | -0.056 (0.026) | 3.25E-02 | -0.001 (0.036) | 0.99 |
| 7-Methylguanine | -0.004 (0.002) | 3.29E-02 | -0.003 (0.003) | 0.34 |
| POV-PC | -0.018 (0.008) | 3.42E-02 | -0.022 (0.011) | 0.05 |
| N-Acetylneuraminate | -0.049 (0.024) | 4.45E-02 | -0.024 (0.034) | 0.47 |
| *Significant in tibia* |  |  |  |  |
| 1-Linoleoylglycerol | 0.020 (0.018) | 0.29 | 0.083 (0.023) | 6.60E-04 |
| Glycochenodeoxycholic acid | 0.009 (0.012) | 0.44 | 0.044 (0.016) | 6.83E-03 |
| Docosahexaenoic acid (DHA) | -0.009 (0.006) | 0.16 | -0.022 (0.008) | 9.21E-03 |
| Allantoin | -0.011 (0.007) | 0.13 | -0.026 (0.010) | 9.56E-03 |
| Glycocholic acid | 0.015 (0.011) | 0.18 | 0.037 (0.015) | 1.74E-02 |
| Malic acid | 0.011 (0.016) | 0.47 | 0.049 (0.021) | 2.07E-02 |
| MEP (Mono-ethyl phthalate) | 0.025 (0.027) | 0.36 | 0.085 (0.037) | 2.32E-02 |
| Oleoyl-Glycerol | -0.011 (0.010) | 0.26 | -0.030 (0.013) | 2.79E-02 |
| L-Isoleucine | 0.007 (0.005) | 0.17 | 0.014 (0.007) | 3.66E-02 |
| Indole-3-Carboxylic acid | 0.004 (0.006) | 0.54 | -0.017 (0.008) | 3.82E-02 |
| Erucic acid | -0.006 (0.006) | 0.32 | -0.017 (0.008) | 3.96E-02 |
| 2-Undecanone | 0.008 (0.005) | 0.07 | 0.013 (0.006) | 4.10E-02 |

Finally, we compared metabolites significantly associated with bone and blood Pb. Two metabolites, glycochenodeoxycholic acid and glycocholic acid, were negatively associated with blood Pb and tibia Pb, while 5-aminopentanoic acid and 7-methylguanine were negatively associated with blood Pb and patella Pb. Further, one metabolite, 5-aminopentanoic acid, was associated with all three measures (**Figure 3**).

**Figure 3.**
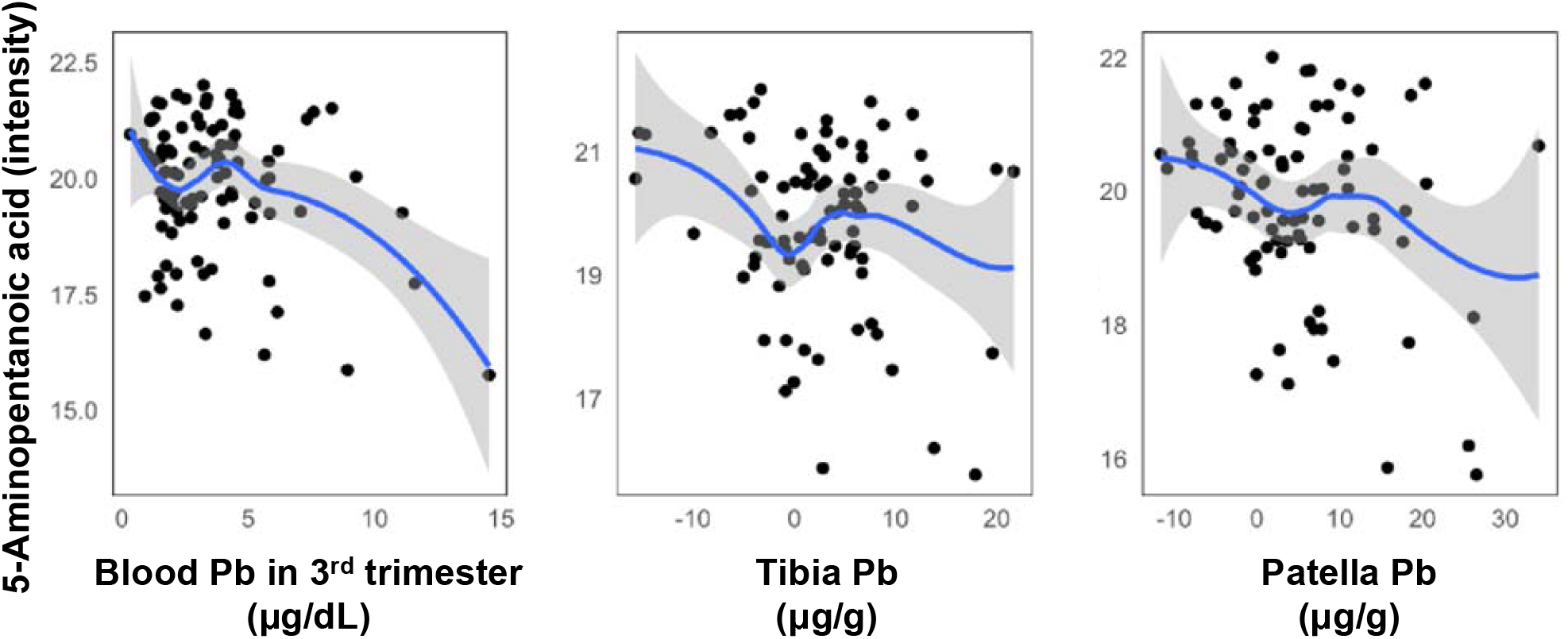
Scatterplots of 5-aminopentanoic acid versus Pb in 3^rd^ trimester blood, tibia, and patella.

## Discussion

Pb exposure is associated with many deleterious effects, but the complex downstream biological perturbations that result from Pb exposure, particularly at lower levels and during pregnancy, are not well understood. Metabolomics, which provides a global, unbiased measurement of circulating metabolites, enables discovery of metabolites and underlying mechanisms associated with exposures. However, to date, population studies on metabolomics of Pb exposures are sparse and have focused on high-exposure populations (12) or have been performed on aging adult men (13). Our study on 99 pregnant women is the first to investigate third-trimester serum metabolomics with both blood Pb and bone Pb contents to unravel biological impacts during pregnancy of both acute and long-term Pb exposure.

Lead exposure through inhalation and ingestion results in absorption into the blood, wherein plasma levels reflect the most rapidly exchangeable fraction of Pb in the bloodstream. This fraction is assumed to be associated with the toxic effects of Pb (27). In addition, around 95% of the total Pb burden is contained within bone, which reflects integrated or cumulative Pb exposure (28). During rapid bone turnover, such as in pregnancy, Pb from bone may migrate into plasma, influencing plasma Pb levels (29). Indeed, we observed a greater number of metabolites associated with blood Pb (31 metabolites) than bone Pb (14 and 11 metabolites in patella and tibia, respectively) in study participants, coinciding with weak to moderate correlations between the three Pb measures (Pearson’s r=0.15-0.51). The third trimester is the period during pregnancy that contains the greatest mobilization of Pb from maternal bone and fastest fetal growth (30,31). Since fetuses are directly exposed to the maternal Pb fraction via umbilical cord transfer (32), higher exposure to Pb during the third trimester may coincide with adverse effects on fetal development. Thus, maternal exposure metrics of Pb are a proxy for fetal exposure, and altered metabolomes that we observed herein may reflect both systemic biological responses (tibia and patella bone Pb) and acute responses (blood Pb) with potential to impact both maternal and fetal health.

Our results show that cumulative Pb exposures measured in bone reflected different metabolite association patterns than acute Pb exposures measured in third-trimester serum (**Tables 2** and **3**). However, two bile acids, glycochenodeoxycholic acid and glycocholic acid, were negatively associated with both blood Pb and tibia Pb. Lead is associated with microbiome dysregulation in animal models (33,34) and in human studies of adults (35) and children (36), and bile acids regulate the gut microbiome as well as host physiology (37). Similar to our findings, Pb exposure in mice for 13 weeks decreased bile acid concentrations measured in stool as well as altered the abundances of bacterial genera, most of which were reduced upon Pb exposure (10). However, this relationship is also likely bidirectional, as the gut microbiome interacts with bile acids (38). In addition, we found a significant negative association between 5-aminopentanoic acid and blood, tibia, and patella Pb. As 5-aminopentanoic acid is a lysine degradation product formed both endogenously and through bacterial catabolism of lysine, these results may further support a non-transient effect of Pb on the human microbiome (39).

We observed that acute Pb exposure (blood Pb) was associated with alterations in saturated and polyunsaturated fatty acids and metabolites in the alpha linoleic acid and linolenic acid pathway. Studies on Pb exposure and lipid alterations in humans are limited. A pilot study of pooled serum samples from occupationally exposed workers to Pb, cadmium, and arsenic found increased levels of very-low-density lipoprotein, decreased low-density lipoprotein, and increased unsaturated fatty acids associated with high heavy metal exposure. These changes in lipid fraction imply disturbance of lipid metabolism (40), though it was not possible to separate the biological effects of individual metals from this mixture. In a single study of 53 postpartum women following delivery, no correlation between fatty acid and Pb levels in maternal blood was observed; notably, that study reported concentrations and ranges of Pb were lower (1.3 [0.6] μg/dL) than in our population of mothers (2.9 [2.6] μg/dL, **Table 1**).

Though human studies of Pb exposure and lipid dysregulation during pregnancy are limited, several studies in rat dams and pups reported associations consistent with our current study. Pb exposure in lactating dams resulted in significantly decreased total monounsaturated fatty acids and total fatty acid concentrations; in particular, reduced levels of dihomo-gamma-linolenic acid (DGLA), linoleic acid (LA), and eicosapentaenoic acid (EPA) were observed in plasma (41). Further, rats exposed to Pb during pregnancy and lactation had reduced DHA in milk and mammary gland tissues (42), and in pups, Pb exposure in dam’s milk, in combination with an *n*-3 insufficient diet, resulted in a reduction in total saturated fatty acids and total *n*-3 and total *n*-6 fatty acids in plasma i (43). Interestingly, the relationship may be bidirectional: a lower blood Pb concentration was observed in rats supplemented with a diet rich in *n*-3 and *n*-6 polyunsaturated fatty acids (44).

Acute Pb exposure was also associated with decreased 2-hydroxybutyrate, a ketone body derived from α-ketobutyrate (alpha-KB) that is produced by amino acid catabolism (threonine and methionine) and glutathione anabolism (cysteine formation pathway) and is metabolized to propionyl-CoA and carbon dioxide (45). 2-hydroxybutyrate levels are hypothesized to be linked with lipid oxidation, oxidative stress, and glutathione synthesis (46). Interestingly, 2-hydroxybutyrate, palmitic acid, palmitoleic acid, stearic, and lactic acid (**Table 2**) are all linked with gestational diabetes during the second trimester and three months postpartum (47), with 2-hydroxybuytrate indicated as an early biomarker of glucose intolerance (48,49). While these metabolites were higher in mothers with gestational diabetes than controls in the previous studies, which is the opposite direction of our findings, Pb levels may be associated with alterations in similar pathways. Indeed, Pb exposure can induce fasting hyperglycemia and glucose intolerance in rats (50). Future mechanistic studies that investigate the role of Pb on pathways involving α-ketobutyrate, such as phenylalanine and tyrosine metabolism, branched chain amino acids metabolism, and homocysteine metabolism, during pregnancy are encouraged.

Organic environmental exposures are linked with blood Pb levels in pregnant women, including caffeine (51). Coffee, as well as tea and coca, contain high levels of Pb (52), possibly due to use of ceramic mugs containing Pb-based glaze (53) or contaminated water used in the coffee-making process and the propensity for Pb absorption into organic matter (54). We observed that serum caffeine and 1-methyluric acid (a major metabolite of caffeine) were positively associated with blood Pb levels, but not bone Pb levels. The association between third-trimester blood Pb and caffeine most likely represents acute exposures since caffeine has an estimated half-life of up to 11 hours during the last weeks of pregnancy (55). Therefore, caffeine, which has been linked with both beneficial effects in adults and adverse fetal health outcomes (56), may be a particularly important Pb co-exposure to consider in future studies.

Betaine was most strongly associated with patella Pb, but also reached marginal significance with blood Pb (p=0.07). Betaine is a vital methyl group donor in transmethylation reactions, as part of one-carbon metabolism via the methionine cycle. Since the availability of methyl groups influences methylation (57), betaine may support a mechanistic premise for the role of one-carbon metabolism in observed associations between blood Pb levels and DNA methylation in highly-exposed populations (58), maternal bone Pb levels and DNA methylation in cord blood (59), and in animal exposure models (60). Interestingly, increased betaine levels were observed in the urine of Pb-exposed workers from a Pb-acid battery recycling site (12), suggesting that increased circulating betaine may be a biomarker of chronic or high Pb exposure levels. Betaine metabolism is also linked to the gut microbiome, specifically probiotic bacterial taxa associated with obesity prevention (*Akkermansia, Bifidobacterium*, and *Lactobacillus*) (61,62). The association between betaine and Pb could be due to Pb-related gut dysbiosis, potentially reducing bacteria that metabolize plant-based molecules.

Our discovery study had several limitations. First, the study included a modest sample size of 99 women in their third trimester of pregnancy from the PROGRESS cohort. While this sample captured an important pregnancy window for Pb exposure, mothers solely resided in the geographic region of Mexico City, which may limit generalizability to other populations. Second, maternal blood was sampled only once during the third trimester and may not reflect fluctuations in Pb levels or metabolite profiles. While we also used bone Pb measurements to investigate associations with cumulative Pb exposure, we cannot rule out that results were affected by sampling time. Third, this study was limited to the investigation of the main effect of Pb, even though environmental exposures to organics, nutritional compounds, and other toxic metals occur simultaneously, with potential synergistic or antagonistic effects on maternal and fetal biology. Finally, given that our study was cross-sectional, causality cannot be addressed.

## Conclusion

This discovery study identified dysregulated metabolite profiles during the third trimester of pregnancy that were associated with blood Pb and bone Pb levels. In particular, altered 2-hydroxybutyrate and metabolites in the alpha linoleic and linolenic acid pathway suggest potential mechanisms linking short-term Pb exposure to adverse maternal health, such as glucose intolerance and gestational diabetes, and a potential effect of Pb on altered gut microbiome and microbial metabolism. Further, Pb exposure levels measured in bone were associated with increased betaine, a methyl donor involved in methylation processes. Together, these results point to several pathways in which short-term and cumulative Pb exposure may affect maternal and fetal health. Although additional replication studies during pregnancy utilizing larger sample numbers and investigating exposure mixtures are encouraged, these results provide insights to guide future detailed mechanistic studies.

## Data Availability

The data that support the findings of this study are available from the PROGRESS study team (PI: Robert O. Wright), but restrictions apply to the availability of these data. Data are available upon reasonable request.

## List of Abbreviations

(AdA): adrenic acid
(ALA): alpha-linolenic acid
(AA): arachidonic acid
(BMI): body mass index
(DGLA): dihomo-gamma-linolenic acid
(DHA): docosahexaenoic acid
(DPA): docosapentaenoic acid
(ENT): effective number of tests
(ETA): eicosatetraenoic acid
(EPA): eicosapentaenoic acid
(GLA): gamma-linolenic acid
(IQR): interquartile range
(Pb): lead
(LA): linoleic acid
(LC-HRMS): liquid chromatography high-resolution mass spectrometry
(Instituto Mexicano del Seguro Social – IMSS): Mexican Social Security System
(‘pooled QC’): pooled quality control sample
(PCs): principal components
(PROGRESS): Programming Research in Obesity, Growth, Environment, and Social Stressors
(RP): reverse phase
(SDA): stearidonic acid

## Declarations

### Ethics approval and consent to participate

Procedures were approved by institutional review boards at the Harvard School of Public Health, Icahn School of Medicine at Mount Sinai, and the Mexican National Institute of Public Health. Women provided written informed consent in Spanish.

### Consent for publication

Not applicable.

### Authors’ contributions

MN and LP contributed to the conception and design of the work, data interpretation, and drafting and revision of the manuscript. SE contributed to the drafting and revision of the manuscript. AJ contributed to the data analysis and revision of the manuscript. GD and CA contributed to the sample analysis, data acquisition, and revision of the manuscript. AC, HL-F, MT-R, RW contributed to the conception and design of the work, data acquisition, and revision of the manuscript. All authors read and approved the final manuscript.

## Acknowledgements

We thank the ABC (American British Cowdray Medical Center) in Mexico for providing research facilities and all of the PROGRESS staff and study participants.

